# Potential Impact of a Diagnostic Test for Detecting Prepatent Guinea Worm Infections in Dogs

**DOI:** 10.1101/2023.10.30.23297718

**Authors:** Hannah Smalley, Pinar Keskinocak, Julie Swann, Christopher Hanna, Adam Weiss

## Abstract

Chad has seen a considerable reduction in cases of Guinea worm disease (or dracunculiasis) in domestic dogs in recent years. Tethering of dogs and application of Abate^®^ larvicide to water sources appear to have contributed to this progress, but with 767 reported dog cases in 2021, accelerating elimination of the disease in Chad may require additional tools. We investigate the potential benefits of a hypothetical diagnostic test which could be capable of detecting pre-patent infections in dogs. We adapt an agent-based simulation model for forecasting the impact of interventions on guinea worm disease in dogs to examine the interaction of multiple test factors including test accuracy, when the test can detect infection, dog selection, and dog-owner compliance with tethering recommendations. We find that a diagnostic test could be successful if used in conjunction with existing interventions, and elimination can be achieved within two years with 80% or higher test sensitivity, 90% or higher specificity, systematic testing of each dog twice per year, and over 90% long-term tethering compliance when a dog tests positive or a worm is emerging. Because of the long incubation period of Guinea worm disease (10-14 months) and the fact that no treatment exists, the benefits of the test rely on the testing rollout and response of dog owners. If the test could estimate the timing of worm emergence, long-term tethering could be eliminated and infected dogs could be tethered only when the worms are expected, minimizing the related resources (human and financial) to support the intervention.

## INTRODUCTION

In 2021, 767 dogs were reportedly infected with Guinea worm disease (or dracunculiasis) in Chad.^1^ This is a vast reduction in cases of Guinea worm (GW) in dogs in recent years, but the number and geographic distribution of cases poses risks for the human population as infections are passed through shared water sources. Chad reported 8 human cases in 2021. There is no treatment for infected animals or humans, and there is no vaccine to prevent infection.^2^

GW is a parasitic nematode. When a dog ingests freshwater copepods harboring infective GW third-stage larvae, either through surface water or a paratenic/transport host, the dog becomes infected and after an incubation period of 10-14 months, a gravid female worm (or worms) emerges from the dog’s skin.^3^ If the emergent worm is exposed to water, tens of thousands of first-stage larvae are released. Copepods ingest the larvae, and the cycle continues.^2^

Interventions employed to reduce dog infections include proactive tethering to minimize exposure to potentially infected water sources, tethering of known infected dogs to prevent the dogs from seeking water sources for relief when worms are emerging, the safe disposal of aquatic waste, and application of Abate^®^ (Ludwigshafen, Germany) larvicide to water sources to kill infectious copepods in the water. These interventions, coupled with robust surveillance, appear to have reduced infections.^4^

When a dog is infected with GW, there are no external signs until near the end of the incubation period.^2^ Based on the experience of field teams in Chad, dogs are often tethered during the day, but some are released at night and fewer are tethered intermittently or not at all. Unfortunately, if a dog is released even just at night, they may still interact with the water sources and aquatic waste and acquire infection or exude worms; “a single missed emerged worm could delay elimination by a year or more.” ^2^ Elimination of the disease may require additional tools to either interrupt transmission or inform the International Commission for the Certification of Dracunculiasis.

In this study, we assess the potential impact of a diagnostic test for GW disease in dogs. If a diagnostic test could detect pre-patent infections in dogs when there are no signs, then tethering resources could be targeted for dogs testing positive. We use an agent-based simulation to model GW infections in dogs over time while accounting for the impacts of currently used interventions as well as a diagnostic test. There are several important test performance factors that need to be considered.^5^ Sensitivity and specificity rates which define the test accuracy represent the probabilities that the test will, in our case, designate an infected dog as positive and an uninfected dog as negative. Levecke et al. (2021) evaluated the minimal and ideal sensitivity and specificity for new diagnostic tests for soil-transmitted helminthiasis (parasitic worms).^6^ Other studies have assessed the sensitivity and specificity requirements for hypothetical diagnostic tests using simulation (e.g., leprosy^7^, statin-induced myopathy^8^, acute lower respiratory infections^9^), including assessing the cost for false positives and false negatives.^8^ Other important factors we consider relate to deployment. Particularly when considering implementation under resource-limited field conditions, performance must be considered in combination with cost and ease of use.^10^ Similar to diagnostic testing for COVID-19, a highly accurate but expensive test that is resource-intensive may not produce desired results in such a setting as quickly as a less accurate but inexpensive and relatively simple test. ^11,12^ Finally, with no treatments available for GW in dogs, we address the necessary response of dog owners to positive test results.

We compare various testing strategies using the simulated number of infections after multiple years of implementing the testing intervention. We determine the necessary test characteristics, deployment features, and owner behaviors for such a test to lead to faster GW elimination than current intervention methods alone.

## DATA AND METHODS

We received data from the Guinea Worm Eradication Program (GWEP) which reports the number of dogs exuding worms and the number of worms exuded by village and month for years 2016-2021. We also collected temperature and rainfall data for the region which is made freely available by the National Oceanic and Atmospheric Administration (NOAA).^13^

We group villages into clusters (“Central”, “East”, and “West”) based on districts along the Chari River (methodology reported by Wang et al.^14^). The number of dogs exuding worms over time by geographic cluster (2016-2021) and by village (year 2021) are reported in Figure 1.

**Figure 1.**
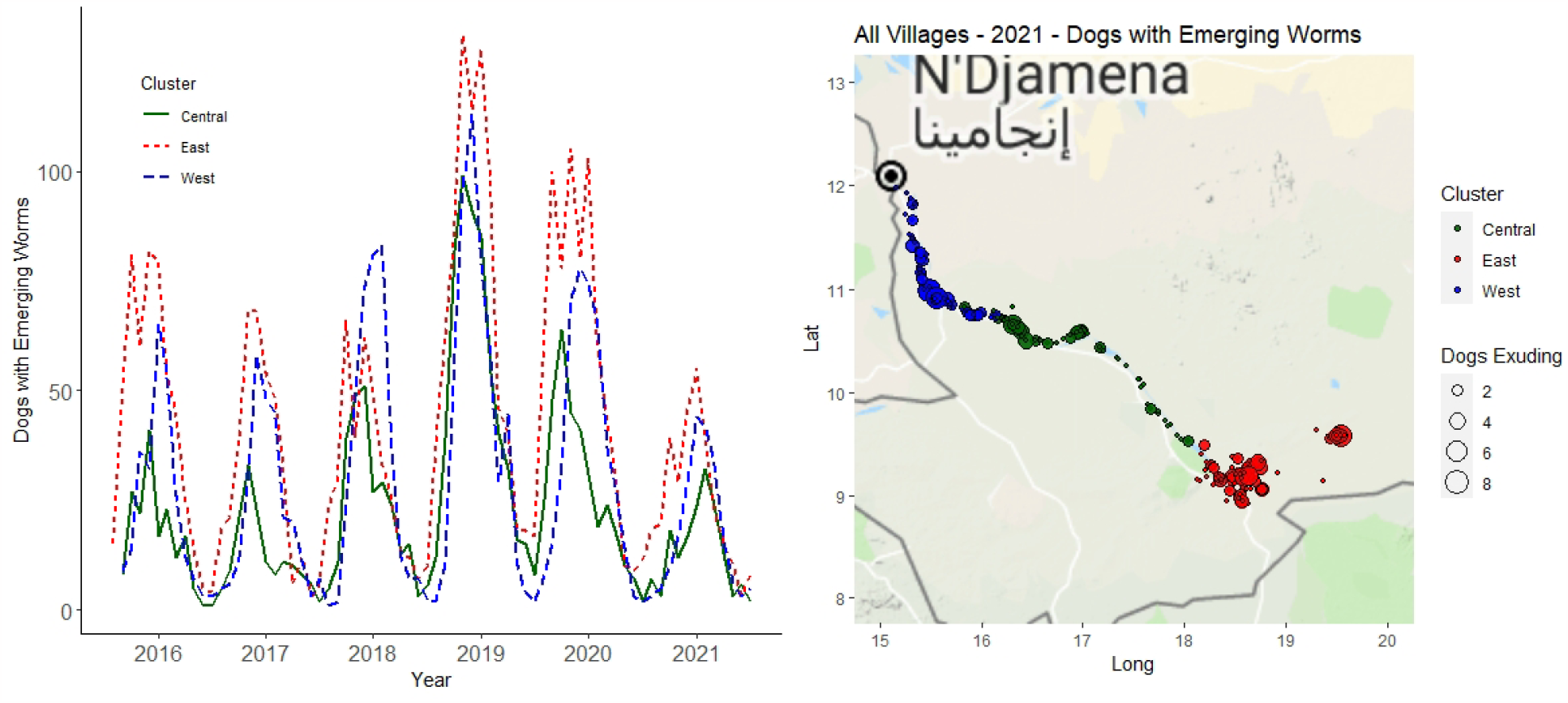
The number of dogs with emerging worms from 2016-2021 by geographic cluster and by village for year 2021.

We adapt the agent-based simulation model developed by Perini et al.^15^ and revised for multiple water sources by Wang et al.^14^ to incorporate the testing intervention. The simulation model simulates daily interactions between dogs, worms, and water sources, considering the life cycle of GW, the impact of Abate^®^ larvicide and weather on water infectivity, and the tethering of dogs. Infectivity and weather parameter values used in this simulation model are reported in the supplemental materials. These parameters were calibrated by Perini et al.^15^ and Wang et al.^14^ based on infection data provided by GWEP and weather data from NOAA for years 2016-2018. In 2019, Chad reported an 86% increase in infected dogs and a 28% increase in villages with infected dogs compared to 2018.^16^ When attempting to recalibrate model parameters using more recent data (i.e., 2019-2021), we find that due to high variability/spikes in dog infections during years 2019 and 2020, we can more closely simulate dog infections in year 2021 by using the original infectivity and weather parameters calibrated based on years 2016-2018 (for details on parameter calibration, see ^14,15^).

During the dry season, dogs may visit multiple water sources but we assume that dogs will only visit their cluster-specific water source during the rainy season because water is more plentiful. Travel behavior for dogs in the dry season and a compartmental diagram of the transmission cycle between dogs and a single water source are presented in the supplemental materials. Dogs become infected by ingesting copepods which carry the infective mature GW larvae or short-term transport/paratenic hosts. A dog with an emerging worm on a given day may infect the water sources it visits. Emerging worms and corresponding GW larvae cohorts which are released into the water are tracked by the simulation; when GW larvae mature (10-14 days), water infectivity increases, and when GW larvae die (after 30 days), water infectivity decreases. Pseudocode for the simulation model is provided in the supplemental materials.

Using the adapted simulation model with the added diagnostic-testing intervention, we examine the interaction and impact of the following factors related to a potential diagnostic test:

1. sensitivity and specificity of the test,
2. timing of when the test can detect infection,
3. which dogs to test (and how often), and
4. dog-owner behaviors (i.e., compliance with tethering recommendations).

Interactions between these factors affect outcomes (e.g., specificity vs. tethering compliance, accuracy vs. reach).^17^

### Sensitivity and Specificity of a Potential GW Diagnostic Test

Test accuracy is determined by the sensitivity and specificity of the test. In our case, the test sensitivity, or true positive rate, is the percentage of infected dogs that will correctly test positive. Specificity, or true negative rate, is the percentage of infection-free dogs that will correctly test negative. We consider varying sensitivity and specificity levels from 60% to 100%. We evaluate the interaction of these values as well as the impacts on potential tethering compliance as the rates of true positives and false positives impact the trust of dog owners and resulting compliance with recommended interventions (i.e., tethering).

### Infection Detection Test Timing

When infection can be detected is an important component in evaluating the potential impact of a diagnostic test. We consider two possibilities:

1. immediate detection: positive result from initial infection until worm emergence, and
2. 30-day detection: worms are detected only if they will emerge within 30 days.

We also consider the possibility that a positive test result under the first option (immediate detection) could include an estimated time interval until worm emergence. This might be possible if the test measures and reports the parasite load and if this value could be used to calculate how long the dog has been infected. Since GW infections in dogs last 10-14 months, testing positive too early during this incubation period without an idea of when to expect worms could undermine tethering compliance.

### Selecting Dogs to Test

We assume a fixed population of dogs. Dogs are assigned to specific clusters but with the potential to travel between clusters based on an ecological study of GW infection in dogs.^18^ We consider two possibilities for dog selection for testing: random and systematic. Under random testing, we consider scenarios with 10% or 20% of dogs randomly selected for testing during each testing period. Under systematic testing, we divide dogs into cohorts, with one cohort tested during each testing period (cohort sizes from 10%-100%). We consider testing dogs monthly throughout the year. Due to the seasonality of GW in Chad, with more instances of worm emergence during the rainy season than the dry season, and because of the long incubation period of GW, we also consider testing dogs only during the off-peak season (dry season) for worm emergence.

### Tethering of Dogs by Owners

Whether or not a dog owner will tether a dog that tests positive for GW is potentially the most important factor in determining the potential impact of a diagnostic test.

Because of the extent of possible owner behaviors and presence of stray or ownerless dogs, we account for various options in the simulation model and make the following assumptions.

▪ <u>When a worm is emerging</u>, we assume there is a 70% probability that the exuding dog will be tethered, which aligns with recent reports.^19,20^
▪ <u>When a dog has a positive test result</u>, we assume that the probability that the dog will be tethered at all is equal to the test specificity level. Scenarios where tethering probability is independent of test specificity lead to unrealistic results. For example, scenarios with high tethering probability and low test specificity lead to better outcomes and faster GW elimination due to high tethering rates of mostly “false positive” dogs. In reality, when specificity is low, dog owners over time will find the test to be unreliable and be less likely to tether dogs following a positive test result.
  ➢ Based on the tethering probability, if a dog is tethered, then they have a daily tethering probability, allowing for the possibility that they could be released at night. We test daily tethering probabilities from 50% to 100%, and consider tethering time periods of 30 days, 90 days, 180 days, and 360 days. If worms emerge from a test-positive (tethered) dog during the tethering time period, we assume 100% tethering and 100% daily tethering probability until the last worm exudes.

We also consider dog owner behaviors for the case where the test will only detect infection within 30 days of worm emergence. In this case, with a positive test result, we assume 100% tethering and 100% daily tethering probability until the last worm exudes. Similarly, if an estimated emergence window could be provided along with a positive test result under the immediate detection test timing scenario, we assume 100% tethering and 100% daily tethering probability during the exuding window, with no change in behavior until then.

### Additional Assumptions

The model includes the following additional assumptions:

- Test results are instantaneous, and the test does not detect past infections.
- Abate^®^ treatment impact = 50%. Bodies of water within a region are treated as a single water source. If Abate^®^ is applied to 50% of the bodies of water, then we assume a 50% loss of infective stage guinea worm larvae in the combined water source.
- No proactive daily tethering apart from positive test results or worm emergence. While proactive daily tethering is an intervention currently employed even in absence of a known infection, we do not include this in the model so that the impact of the testing intervention and tethering response can be more readily related to reductions in simulated infections.

Figure 2 provides a conceptual diagram of monthly dog diagnostic testing.

**Figure 2.**
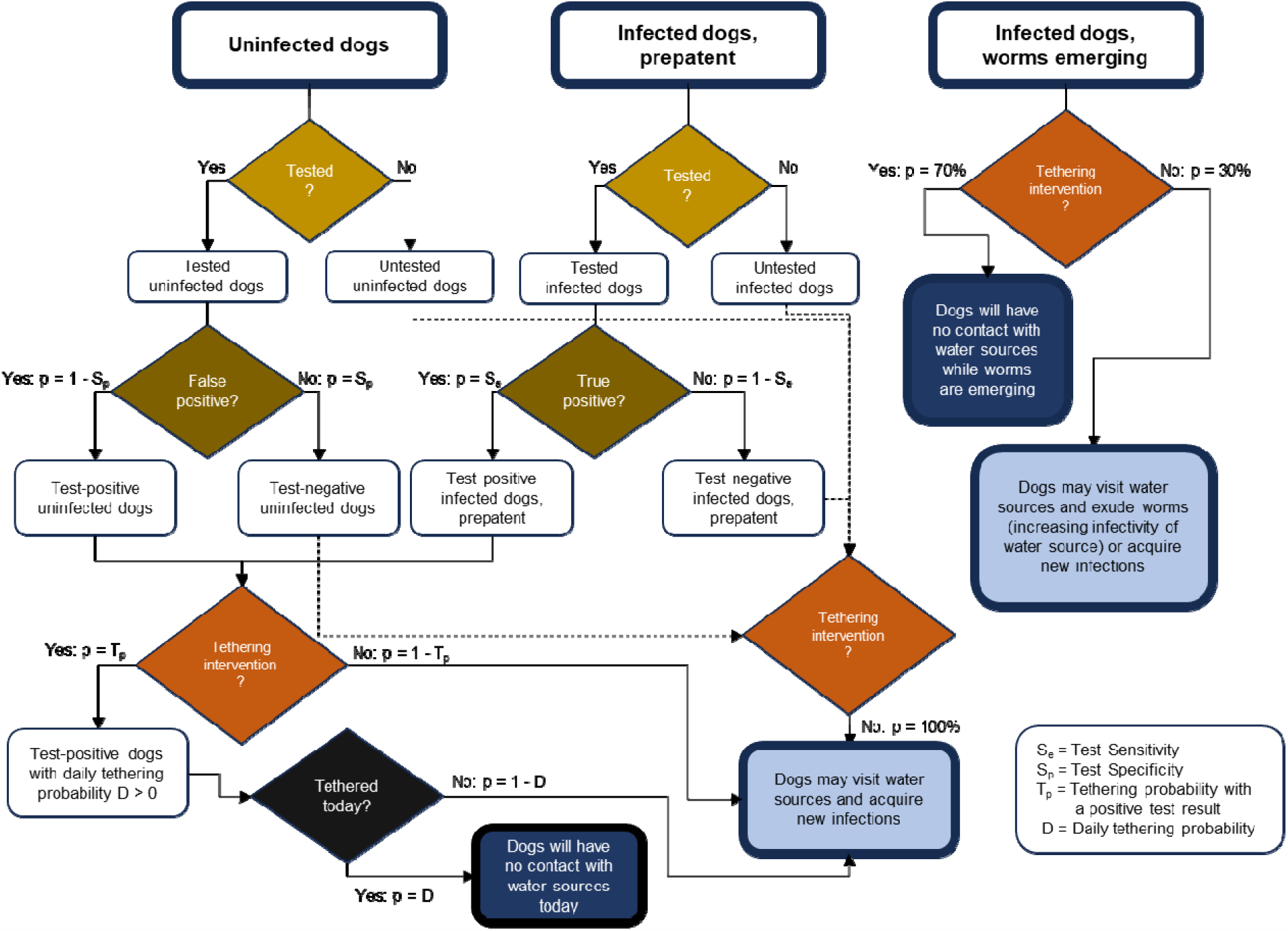
Conceptual diagram of monthly dog testing.

We report results based on the average over 50 simulation iterations for each scenario; notably, we observe minimal variation in results as iterations increase above 20.

## RESULTS

### The impact of random versus systematic selection of dogs for testing

Selecting dogs for testing systematically is more beneficial than selecting dogs randomly; systematic selection helps reduce the number of repeat tests given to a dog within a one-year timeframe. Table 1 reports the percentage of dogs infected after 5 years of intervention for different scenarios with respect to dog selection (random vs. systematic) as well as other factors including the percentage of dogs tested monthly, test sensitivity, and owner behaviors (daily tethering probability and length of tethering). For these scenarios, we assume 100% test specificity and immediate infection detection. If the dogs tested each month are selected randomly, then one year of tethering following a positive test result far outperforms 30-day tethering, but elimination is not achieved within 5 years for either case. If dogs are selected systematically and tethered for one year following a positive result, then elimination can be achieved if 10% of dogs are tested monthly and test sensitivity is 100%, or if 20% of dogs are tested monthly with 90% or higher test sensitivity. As test sensitivity goes down, infections increase but only under 1-year tethering. 30-day tethering is insufficient to reduce infections over time regardless of test sensitivity.

**Table 1.** The percentage of dogs infected after 5 years of the testing intervention under different scenarios with respect to daily tethering probability, length of time for daily tethering, sensitivity, dog selection (random vs. systematic), and percentage of dogs tested monthly.

|  |  | 30-Day Tethering |  | 1-Year Tethering |  | 1-Year Tethering |  |
| --- | --- | --- | --- | --- | --- | --- | --- |
| Daily Tethering | (Se, Sp) | Dog Selection: Random |  | Dog Selection: Random |  | Dog Selection: Systematic |  |
|  |  | 20% | 10% | 20% | 10% | 20% | 10% |
| 50% | (1,1) | 1.54% | 1.56% | 0.35% | 1.35% | 0.00% | 0.00% |
|  | (.9,1) | 1.54% | 1.55% | 0.55% | 1.38% | 0.00% | 0.32% |
|  | (.8,1) | 1.54% | 1.56% | 0.81% | 1.41% | 0.02% | 0.98% |
|  | (.7,1) | 1.55% | 1.55% | 1.04% | 1.43% | 0.22% | 1.28% |
| 70% | (1,1) | 1.53% | 1.54% | 0.27% | 1.31% | 0.00% | 0.00% |
|  | (.9,1) | 1.54% | 1.55% | 0.54% | 1.35% | 0.00% | 0.25% |
|  | (.8,1) | 1.53% | 1.54% | 0.76% | 1.39% | 0.01% | 1.00% |
|  | (.7,1) | 1.55% | 1.55% | 1.05% | 1.43% | 0.25% | 1.24% |
| 90% | (1,1) | 1.53% | 1.55% | 0.36% | 1.30% | 0.00% | 0.00% |
|  | (.9,1) | 1.55% | 1.54% | 0.55% | 1.35% | 0.00% | 0.29% |
|  | (.8,1) | 1.55% | 1.55% | 0.78% | 1.40% | 0.02% | 0.97% |
|  | (.7,1) | 1.53% | 1.54% | 0.99% | 1.44% | 0.25% | 1.26% |

### The interactions between test accuracy, tethering, and the percentage of dogs tested

Figure 3 reports the percentage of dogs infected after 5 years for systematic dog selection and varying test sensitivities and specificities from 60%-100%, tethering time periods (90, 180, and 360 days), daily tethering probabilities (50%, 70%, and 90%) and percentage of dogs tested monthly (cohort sizes from 10%-100%); we assume immediate infection detection. As either the tethering time period increases in length, the percentage of dogs tested monthly increases, or test sensitivity increases, the percentage of infections is reduced. In some cases, lower test specificity results in a lower percentage of infections due to the higher number of dogs tethered because of increases in false positive test results. In general, however, higher test specificity aligns with reductions in infections. The percentage of dogs infected after 5 years when also considering varying Abate^®^ impact levels are reported in the supplemental material.

**Figure 3.**
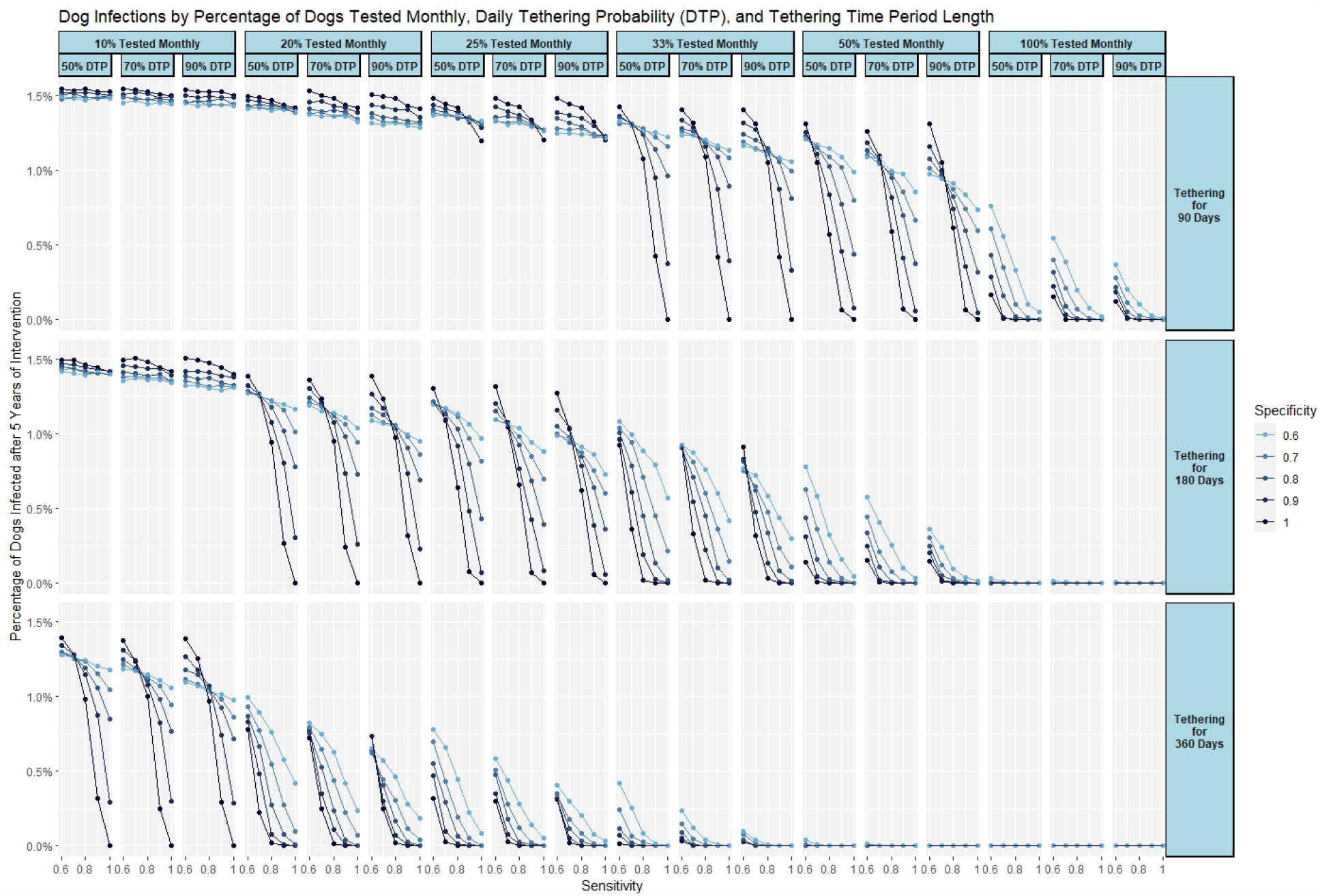
Percentage of dog infections after 5 years of the testing intervention, by daily tethering probability and length of tethering, test cohort size (systematic testing), sensitivity, and specificity.

### Impact of estimated worm emergence on tethering requirements

Figure 4 reports the expected year of Guinea worm elimination (assuming the testing intervention began in 2022) and the total number of dogs that would need to be tethered monthly for 2 scenarios: (i) similar to the analysis above, the test will return only a positive or negative (immediate detection), and (ii) the test will return a positive or negative but with the addition of an estimated worm emergence window. We assume 100% sensitivity and specificity for these scenarios. Elimination can be achieved in 2 years with 1-year tethering and 10% of dogs tested monthly. With the addition of worm emergence estimates, elimination can be achieved in 2 years with 20% or more dogs tested monthly but fewer dogs tethered and for shorter time periods. If dogs are only tested during the dry season, testing 25% monthly can lead to elimination in 2 years.

**Figure 4.**
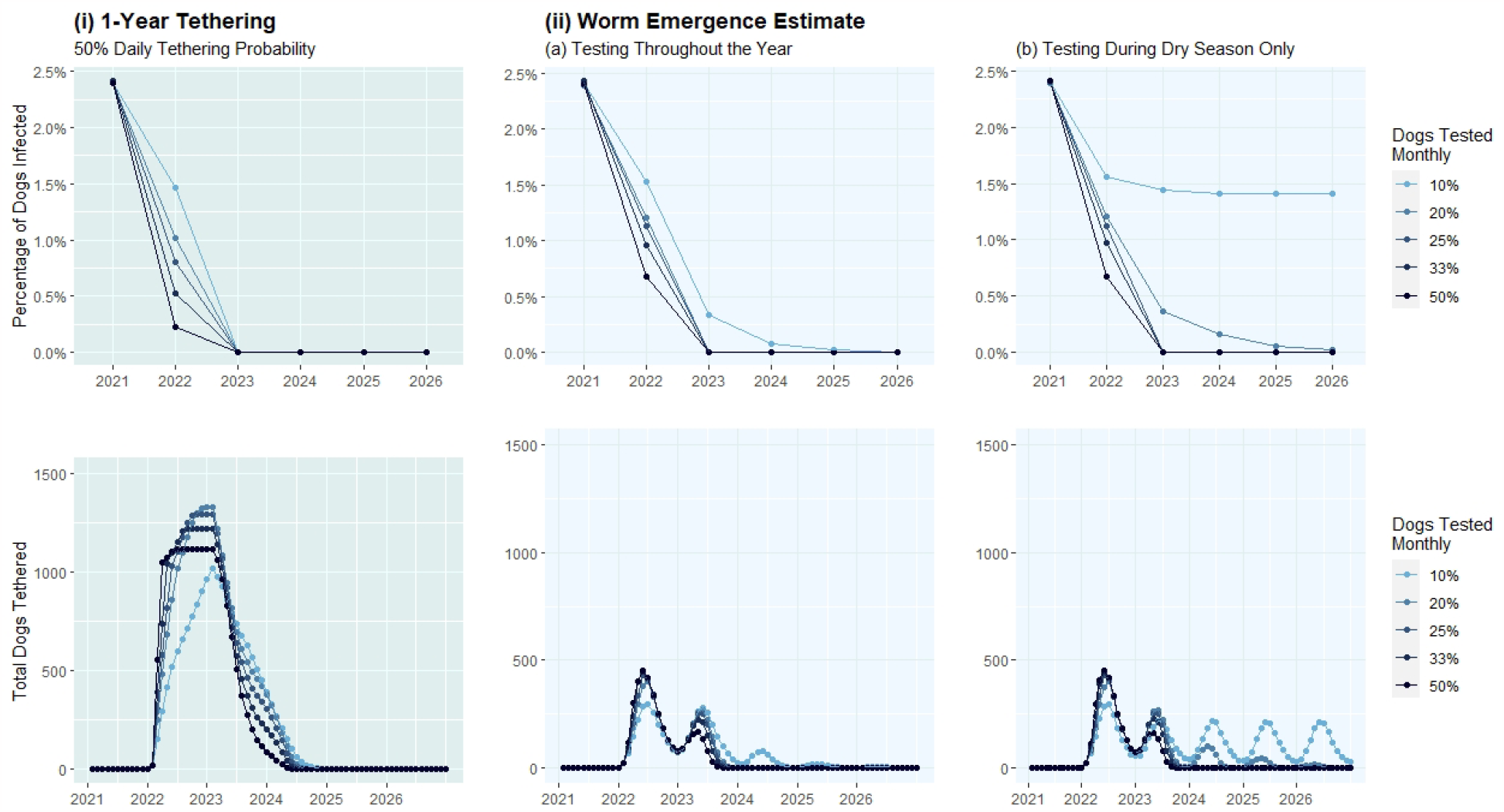
The percentage of dog infections after five years of intervention and total dogs tethered monthly under two testing strategies: (i) “positive/negative only” test and 50% daily tethering probability for one year following a positive test result, (ii) test which reports positive/negative as well as an estimated worm emergence window with tethering only during the test-positive dog’s worm emergence window, with (a) testing throughout the year and (b) testing only during the dry season.

### 30-day infection detection

If infections are detectable only within 30 days prior to worm emergence, then we find that more than 50% of dogs must be tested monthly to see a long-term reduction in dog infections (dogs selected randomly or systematically). Elimination can be achieved within 5 years for this scenario with a perfect test (100% sensitivity and specificity) and 100% tethering compliance only if 100% of dogs are tested monthly.

## DISCUSSION

We find that a diagnostic test could be successful if used in conjunction with existing interventions, and elimination can be achieved under the following conditions:

- 80% test sensitivity, 90% test specificity
- Systematic testing of each dog twice per year
- Long-term tethering (360 days) when a dog tests positive
- Very high tethering compliance (over 90%) when a dog tests positive or a worm is emerging.

Systematically selecting dogs for testing is more effective than selecting dogs randomly. Fewer dogs could be tested monthly when dogs are selected systematically; testing 10% of dogs monthly with dogs chosen systematically results in fewer infections than testing 20% of dogs monthly with dogs chosen randomly, for test sensitivities of 90% or higher (Table 1).

Shorter tethering time periods require higher test sensitivities and more frequent testing to lead to elimination. Based on Figure 3, if the test is perfect (100% sensitivity, 100% specificity), then elimination can be achieved within five years under the following owner tethering behaviors and corresponding percentages of dogs tested monthly:

▪ 360-day tethering → 10% of dogs tested monthly
▪ 180-day tethering → 20% of dogs tested monthly
▪ 90-day tethering → 33% of dogs tested monthly

As sensitivity and specificity decrease, the necessary tethering time to achieve elimination increases. Notably, the impact of the test is heavily dependent on the length of tethering following a positive test result, regardless of the sensitivity and specificity of the test.

The reported scenarios assume 50% Abate^®^ impact and no proactive tethering. Proactive tethering is an important tool for reducing the spread of GW. However, by assuming no proactive tethering in the model, we can more confidently label greater reductions in GW disease over time as direct consequences of the proposed testing intervention. Given the use of proactive tethering in the field, the number of infected dogs reported herein should be overestimations and elimination could be achieved even faster.

Guinea worm infections among dogs has low prevalence when considering the total population of dogs; less than 3% of dogs were reported to be infected in 2021, based on data provided by the GWEP. Thus, short of 100% test specificity, testing in a large but low prevalence population will result in many false positives. Because there is no treatment for guinea worm disease, the response to a positive test result is tethering of the infected dog and requires continued and long-term efforts by the dog’s owner. Over time, if a dog tests positive and is therefore tethered but no worms emerge, owners may begin to doubt the test results and no longer comply with tethering recommendations. A high rate of false positives will undermine the goals of diagnostic testing for GW infections because dog owner behaviors are essential to elimination.

The requirement for long-term tethering could be eliminated if the test could provide an estimate of the timing of worm emergence. Tethering for 1 year following a positive test result, with a daily tethering probability of 50%, can achieve elimination within 2 years even with only 10% of dogs tested monthly (for 100% test sensitivity and specificity). However, this result relies heavily on tethering consistency over a long period of time, requiring tethering of more than 1,000 dogs at one time in response to testing and/or worm emergence (Figure 4). If the test provided an estimate of the timing of worm emergence, test-positive dogs would only need to be tethered when the worms are expected; elimination could be achieved and the monthly tethering burden would be considerably lower. Monthly testing of 10% of dogs throughout the year is sufficient to achieve elimination in less than 4 years under this scenario. Increasing testing capacity to 20% of dogs monthly could achieve elimination in 2 years, with a higher burden on testing resources but without the reliance on long-term tethering compliance. Testing dogs only during the dry season produces similar results if testing capacities of 25% of dogs or more each month can be achieved. Concentrated high-capacity testing efforts in the dry season could reduce (or eliminate) the need for testing in the wet season, when efforts could be focused towards tethering (with some crossover of tethering into the dry season).

If the test could only detect infection within 30 days of worm emergence, elimination can only be achieved within five years if 100% of dogs are tested monthly and test-positive dogs are tethered until all worms emerge. This is unlikely to be feasible. Even testing the same dogs as frequently as every other month will not lead to elimination. If infection could be detected 60 days from worm emergence, with test-positive dogs tethered until all worms emerge, then testing dogs every other month would indeed lead to elimination but at a high testing burden.

Depending on owner behaviors, testing for prepatent infections could have negative consequences for elimination. Owners may choose to untether test-negative dogs; untethered dogs could be exposed to infections in the water sources. Testing too soon after initial infection without any knowledge of worm emergence timing could also have negative consequences. If a dog tests positive but no worm emerges within a few months, then the owner may untether the dog, possibly due to a loss of confidence in the test or simply fatigue from implementing the intervention for so long. It is possible that the test could accurately detect a GW infection (true positive) but a worm may not emerge (e.g., a worm may not reach the surface of the skin or the detected worm could be male^21^). Trust in the accuracy of the test could fall as these cases could be viewed as false positives. Because of the long incubation period and the fact that no treatment exists, the rollout of the test and the response to a positive test result are critical for a test to lead to faster elimination.

### Limitations

In practice, test sensitivity may be higher than expected if test-positive dogs which do not exude worms are assumed to be uninfected rather than possible true-positive cases without worm emergence. There may be logistical challenges to following up with specific dogs (systematic testing) as microchipping and/or unique identifiers are not perfect and Chad experiences high dog mortality. The reported average lifespan of a dog living along the Chari River in Chad is 3 years. Our model is flexible to incorporate age and corresponding death probabilities but does not currently take into account dog births or deaths; rather, the model maintains the same group of dogs throughout the simulation. Data is not currently available which accurately tracks dog populations over time so this is left to future research. However, the probabilistic nature of the model allows us to capture other heterogeneities in the dog population (e.g., behaviors, susceptibility).

We assume 44,000 dogs living along the Chari River based on regional data reported for years 2016-2018, and infectivity parameters were calibrated based on this population size. However, recent estimates report that the population size for this region may be closer to 28,000. We repeated the same analyses reported in Figure 3 with this smaller dog population size (keeping infectivity parameters the same), and the results (percent of infections) are similar and comparisons between different intervention strategies (e.g., tethering timing, dog selection) still hold. The model does not consider diminishing tethering probabilities over time for test specificities below 100%.

A diagnostic test could be a great asset in the fight for GW elimination. However, given that there is no existing vaccine or treatment and because of the long incubation period of GW, the impact of the test will greatly depend on the response of owners when a dog tests positive. The strategy used to implement the test (i.e., dog selection and frequency) and education about the importance of tethering compliance are critical factors to success.

This work has supported the development of a Target Product Profile (TPP) for a diagnostic test to detect Guinea worm infection in animals. The TPP draft was released to the public for comment by the World Health Organization on Oct. 6, 2023.^22^

## Data Availability

All data produced in the present study are available upon reasonable request to the authors.

## Acknowledgements

We thank Tyler Perini, Yifan Wang, Fernando Torres and Tchindebet Ouakou. This study was supported by a grant from the Carter Center; the Global Campaign to Eradicate Dracunculiasis receives financial support from a large coalition of organizations and agencies. Please refer to: www.cartercenter.org/donate/corporate-government-foundation-partners/index.html. This research was also supported in part by the Center for Health and Humanitarian Systems and the following Georgia Tech benefactors: William W. George, Andrea Laliberte, Joseph C. Mello, Richard “Rick” E. and Charlene Zalesky.

