## Supplemental methods and figures for "Potential Impact of a Diagnostic Test for Detecting Prepatent Guinea Worm Infections in Dogs"

##### Section 1: Travel Behavior of Dogs

Each dog is assigned to a subgroup in their home geographic cluster, with each subgroup corresponding to the likelihood of travel to each of the three water sources; dogs do not change subgroups. All dogs use their home water source exclusively during the rainy season (from June to October). The probability that a dog travels to a geographic cluster's water source during the dry season is outlined in Table 1. These probabilities were determined based on research which suggests that a dog's mean travel range is 4.4 $km^2$ in Chad during the dry season, and 80% of dogs visit ponds within 100 meters from the dog owner's home (McDonald et al. 2020).

*Table S1 The probability that dogs in each cluster will visit the different water sources during the dry season.*

| | | West Water Source | Central Water Source | East Water Source |
|---|---|---|---|---|
| West Dogs | Group I: 60% | 100% | | |
| | Group II: 40% | 80% | 20% | |
| Central Dogs | Group I: 40% | 20% | 80% | |
| | Group II: 50% | | 100% | |
| | Group III: 10% | | 80% | 20% |
| East Dogs | Group I: 10% | | 20% | 80% |
| | Group II: 90% | | | 100% |

#### Section 2: Model of Water Infectivity

The Chad GW Eradication Program (GWEP) documented GW infections in Chad in 19 districts representing 1674 villages. Wang et al. used K-means to group districts into three geographic clusters/regions using relative worm emergence per month, latitude, longitude, position along the Chari river and elevation.

Given parameters reported in Table S2, the probability that a dog is infected after visiting a water source on a given day (by consuming infectious copepods via drinking water or transport hosts) is calculated as follows. With the sigmoidal function given by (1), the basic infectivity of a water source when there are n infectious guinea worms in the water source is given by (2). With the addition of seasonality (with environmental factors developed by Perini et al. and based on rainfall and temperature measurements to account for dry vs. rainy seasons), the final infectivity function is given by (3). The infectivity parameters (Table S2) were calibrated by Wang et al. A compartmental diagram of the transmission cycle between dogs and a single water source is presented in Figure S1.

*Table S2 Parameters for water infectivity calculations.*

| | | **Region** | | |
|---|---|---|---|---|
| | **Description** | West | Central | East |
| $F_L$ | Minimum rate of infection | 3.4e-04 | 1.55e-04 | 1.53e-04 |
| $F_U$ | Maximum rate of infection | 4.265e-03 | 3.983e-03 | 2.472e-03 |
| D | Inflection point | 78 | 88 | 125 |
| C | Curvature | 0.0668 | 0.1210 | 0.1661 |
| $w_m$ | Environmental factor in month m: [0.0981, 0.2481, 0.2069, 0.9669, 0.9984, 1.00, 0.4977, 0.5968, 0.3446, 0.1808, 0.2520, 0.1018] | | | |
| $n$ | Number of infectious guinea worms in the water source (worm burden) | | | |
| $a$ | Abate level | | | |

(1) $$f(n) = \frac{1}{1 + e^{-C(n-D)}}$$

(2) $$F(n) = [f(n) - f(0)](F_U - F_L) + F_L$$

(3) $$F_{inf}(n, m) = w_m \cdot (1 - a) \cdot F(n)$$

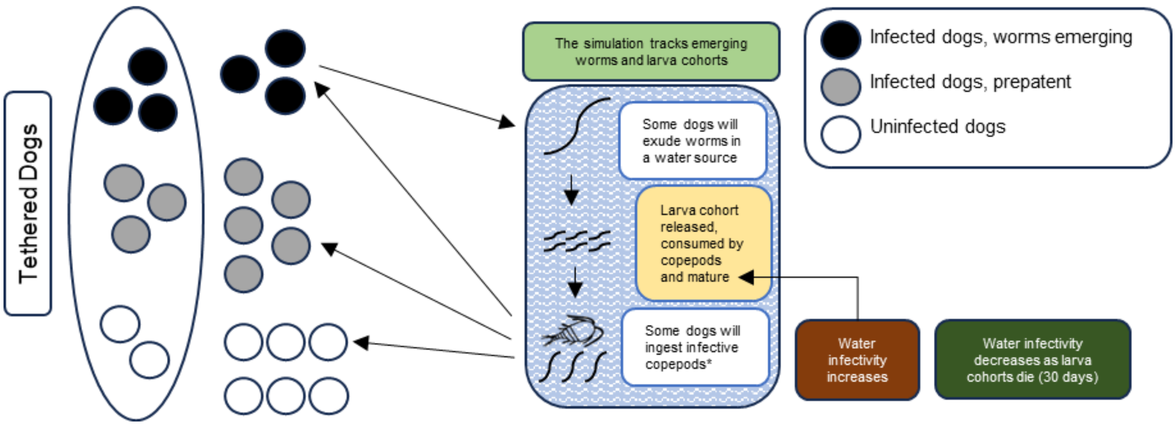


*Figure S1 Diagram of the transmission cycle between dogs and a single water source on a given day, and the impact of exuded worms on the infectivity of a water source. In the dry season, dogs can travel to multiple water sources. If a dog is tethered, it does not visit a water source. Overall water infectivity is determined by environmental factors, abate levels, infectivity parameters, and the number of mature guinea worms in the water. *Dogs are infected by ingesting infective copepods via consumption of water containing copepods or by consuming short-term transport/paratenic hosts.*

### Section 3: Simulation Model with Testing Intervention

#### **Algorithm 1** Pseudo code for the simulation

**Set parameters:**

$nbDogs_1$, $nbDogs_2$, $nbDogs_3$ = set of dogs in regions 1, 2, and 3 of lengths $N_1$, $N_2$, and $N_3$, respectively

$T$ = probability that a dog with emerging worms will be tethered

$A$ = Abate level

$T_{days}$ = length of tethering following a positive test result

$Se$, $Sp$ = test sensitivity and specificity, respectively

$P$ = percentage of dog population to test monthly

$D_{tp}$ = daily tethering probability for test-positive dogs that are partially tethered

**Define**:

$DW(i,r)$ : water contact probability for each dog $i$ and water source/region $r$ (based on travel behavior)

*randvec* = vector of length N of random values (uniform distribution) in (0,1)

**for** iteration 1:20

*#Initialization*

**Set** number of dogs infected, worms exuded per region in previous year (empirical data)

**Assign** dogs proportionally by region to one of $\frac{N}{P \cdot N}$ cohorts (total cohort size = $P \cdot N$)

$\boldsymbol{C}$ = 1 (begin with cohort 1)

**Choose** sample of dogs per region to be infected (random based on empirical data)

**Assign** worms per dog (worms per dog distribution), **calculate** emergence day per worm

**for** day $d$ from 1 to 360*(1 year initialization + 5 years forecasting)

**Define (**per region $r$) the count of infectious worms in the water =

previous count of worms – worms dying on day $d$

\+ worms maturing to infectiousness on day $d$

**if** day $d$ is the first day of the month (($d$ – 1) MOD 30 = 0) and $d$ > 360 (after initialization year) **then**

*#Identify dogs targeted with testing intervention*

$T_{Udogs}$, $T_{Idogs}$ = set of uninfected and infected dogs in cohort$\{C\}$, respectively

*#Determine test-positive dogs and partially-tethered dogs*

$Test_{pos}$ = [random sample from $T_{Idogs}$, length = ceil ($Se \cdot |T_{Idogs}|$ )] ∪

[random sample from $T_{Udogs}$, length = ceil ((1-$Sp$) $\cdot |T_{Udogs}|$)]

$Test_{pos\_t}$ (set of partially-tethered dogs in $Test_{pos}$ ) =

[random sample from $Test_{pos}$, length = ceil (Sp $\cdot |Test_{pos}|$)]

**Update** list of dogs partially tethered due to positive test result

**Assign** end-tethering date for dogs in $Test_{pos\_t}$ = $d$ + $T_{days}$

$C$ = $C$+1

**end if**

*#Determine dogs tethered due to emerging worms*

**for** each region $r$:

**Identify** dogs with emerging worms on day $d$

**for** each dog with emerging worms on day $d$

**if** generated random value in (0,1) ≤ $T$ **or** dog tested positive within $T_{days}$ days **then**

dog is tethered for 30 days

**end if**

**end for**

**Update** count of dogs with emerging worms and number of worms exuded

**end for**

*#Calculate water source infectivity*

**for** each region *r*:

**if** current month is in dry season, **then** new Guinea worms in the water =

the count of emerging worms on day *d* from *non-tethered* dogs with $DW(i,r) > 10\%$

**else,** = the count of emerging worms on day *d* from *non-tethered* dogs from region *r*.

**end if**

**Update** count of guinea worms in the water in region *r*

**Update** lists of dates of when worms are expected to reach infectiousness and death

*#Determine impact of Abate*

**Define** Guinea worms killed *k* = random number (binomial distribution: $n$ = number worms in the water in region *r*, $p = A$).

**Subtract** *k* infectious worms (random) from the list of worms in the water in region *r*

*#Calculate water source infectivity:*

**Calculate** $W(r)$ **=** water infectivity rate based on environmental factor, abate level, infectivity parameters, and count of Guinea worms in the water

**end for**

*#Identify dogs that are infected by a water source on day d*

**for** each region *r***:**

**Define** threshold vector *V* for dogs from *r*:

**If** current month in dry season, **then** dogs may visit alternative water sources:

$V = DW(:,1) \cdot W(1) + DW(:,2) \cdot W(2) + DW(:,3) \cdot W(3)$

**otherwise,** $V = DW(:,r) \cdot W(r)$.

**end if**

**for** each test-positive partially-tethered dog *i*

dog *i* is tethered today if generated random value in $(0,1) \leq D_{tp}$

**end for**

**If** $randvec(i) < V(i)$ (*non-tethered dogs only*), **then** dog *i* is infected on day *d*

**Assign** worms per dog (worms per dog distribution)

**Calculate** worm emergence days

**end if**

**Update** lists of infected dogs and worm emergence days

**end for**

**end for**

**end for**

**return** mean across iterations of worms exuded and dogs with emerging worms by region/day

#### Section 4: Varying Abate Levels

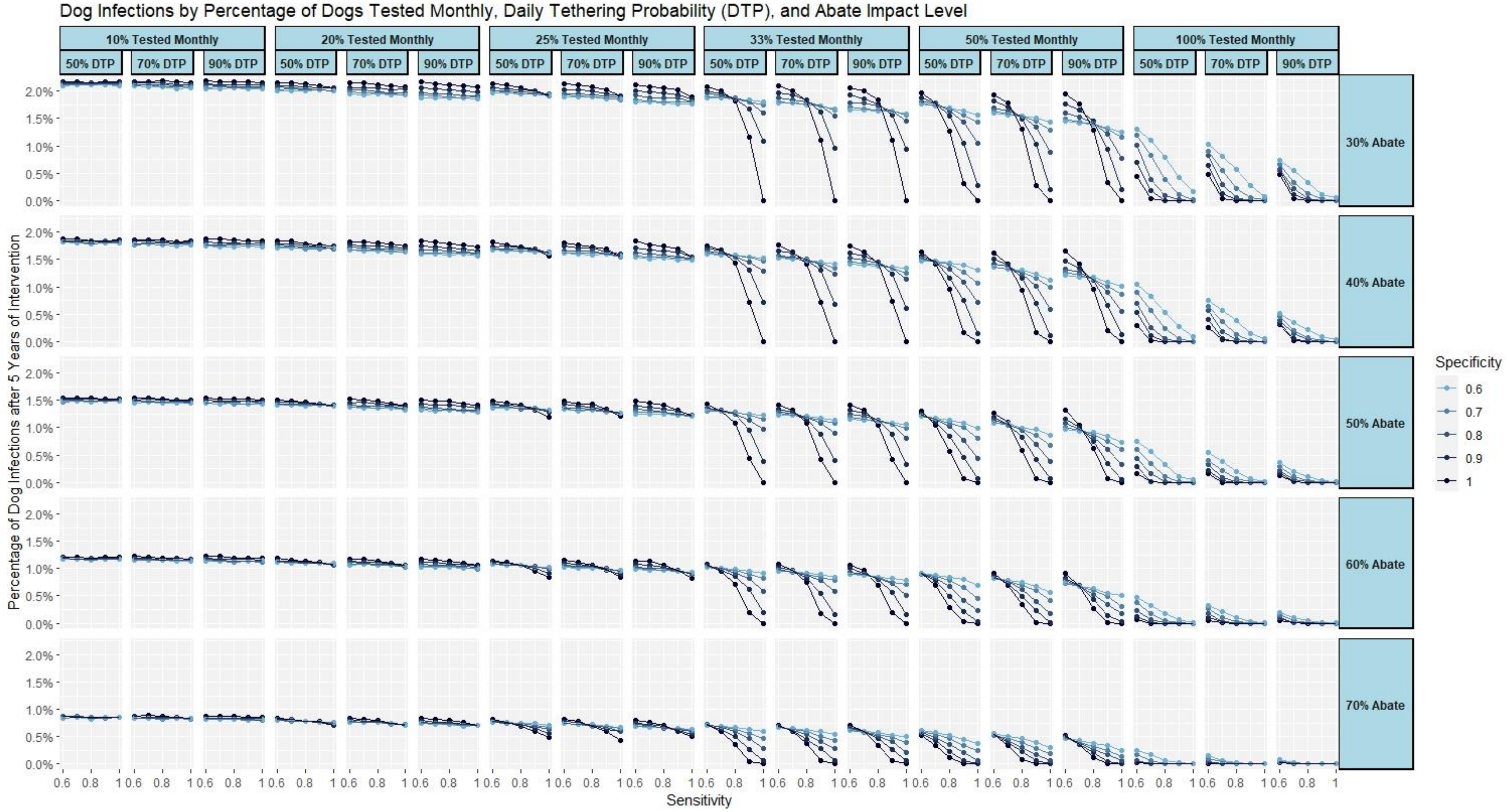


*Figure S2 Percentage of dog infections after 5 years of the testing intervention for 90-day tethering, by daily tethering probability and abate impact level, test cohort size (systematic testing), sensitivity, and specificity.*

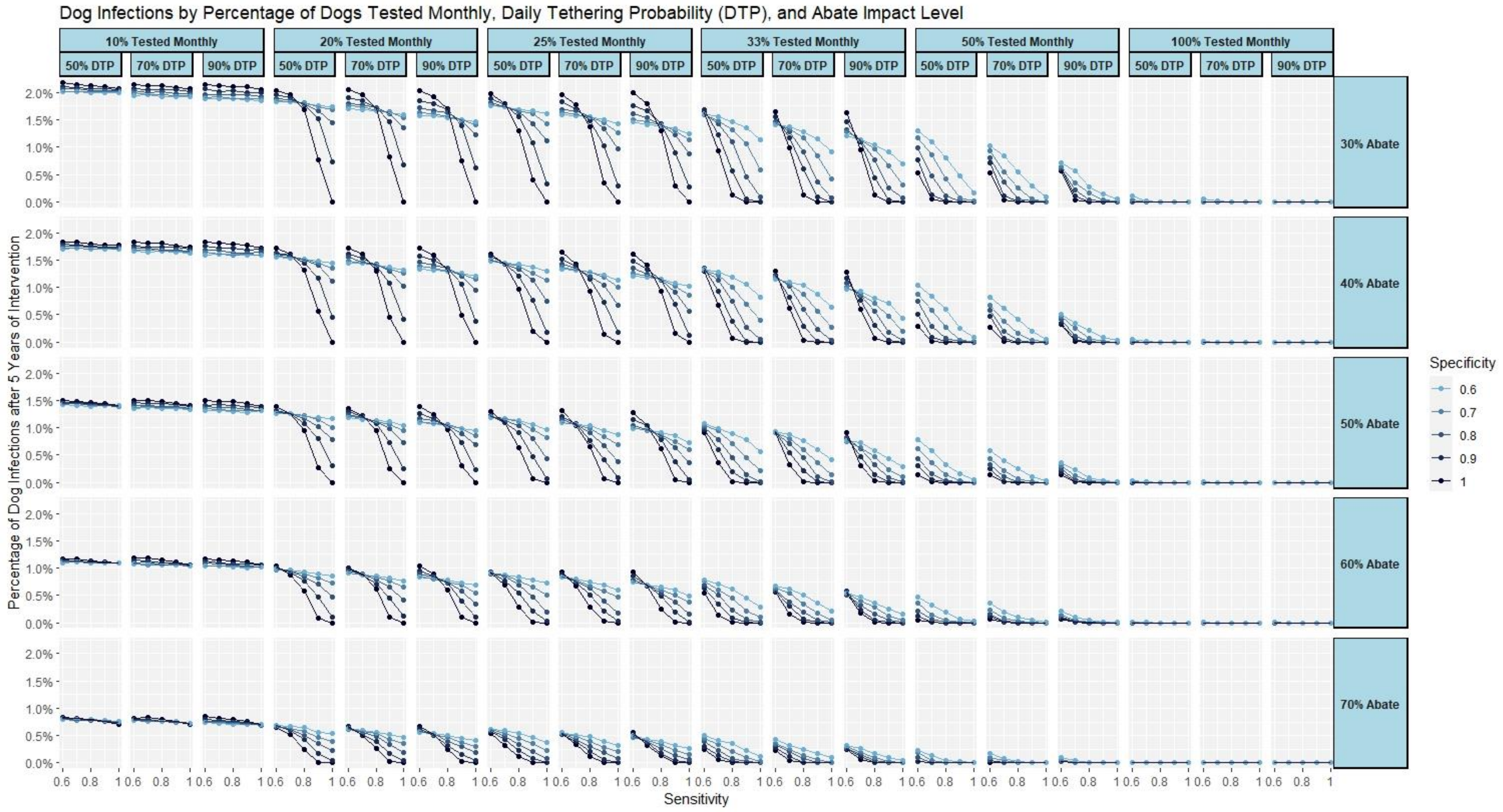


*Figure S3 Percentage of dog infections after 5 years of the testing intervention for 180-day tethering, by daily tethering probability and abate impact level, test cohort size (systematic testing), sensitivity, and specificity.*

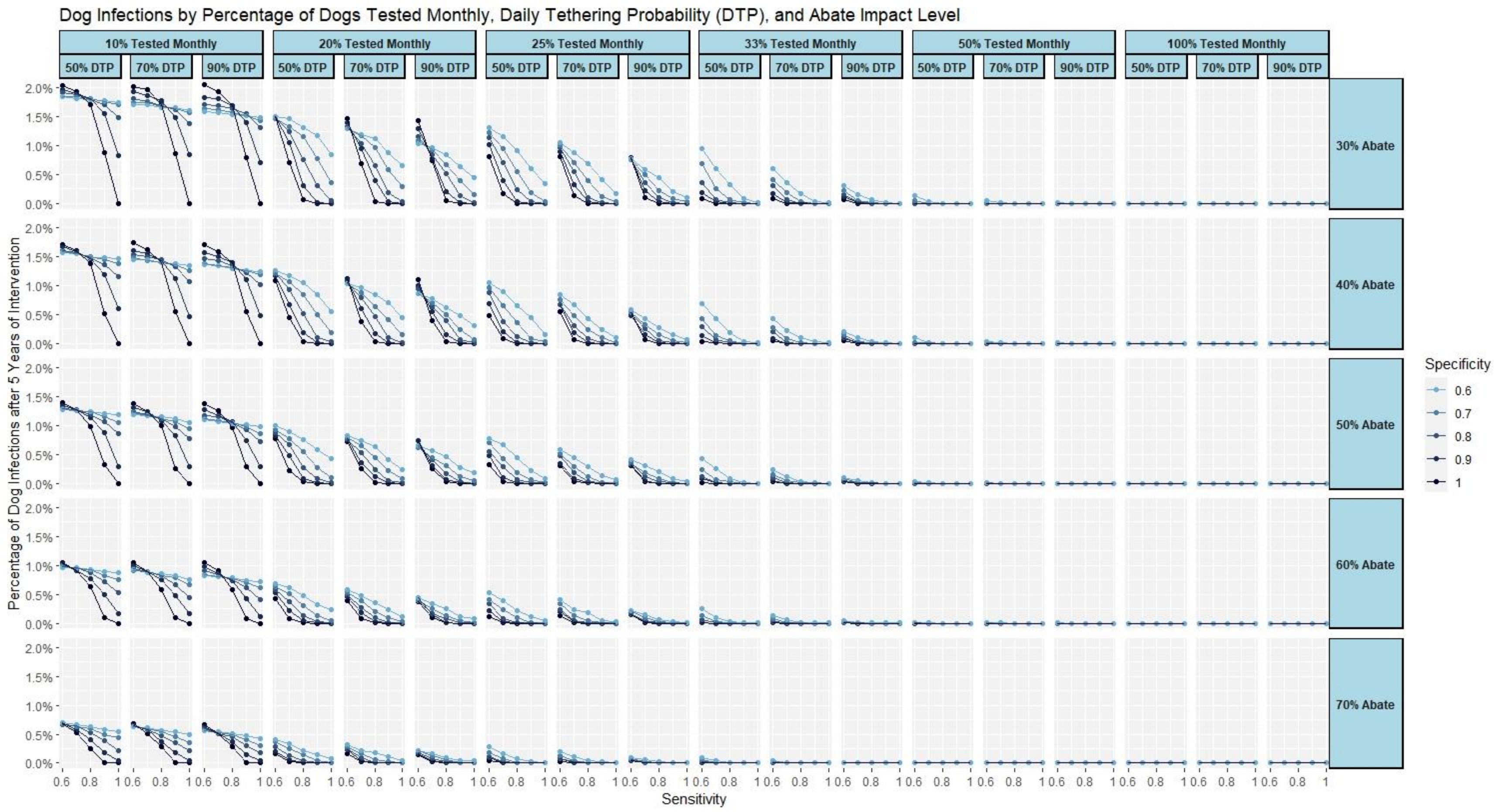


*Figure S4 Percentage of dog infections after 5 years of the testing intervention for 360-day tethering, by daily tethering probability and abate impact level, test cohort size (systematic testing), sensitivity, and specificity.*
